# Hydroxychloroquine inhibits proteolytic processing of endogenous TLR7 protein in human primary plasmacytoid dendritic cells

**DOI:** 10.1101/2021.09.14.21263569

**Authors:** Claire Cenac, Mariette Ducatez, Jean-Charles Guéry

## Abstract

Toll-like receptor 7 (TLR7) triggers antiviral immune responses through its capacity to recognize single-stranded RNA. Proteolytic cleavage of TLR7 protein is required for its functional maturation in the endosomal compartment. Structural studies demonstrated that the N- and C-terminal domains of TLR7 are connected and involved in ligand binding after cleavage. Hydroxychloroquine (HCQ), an antimalarial drug, has been studied for its antiviral effects. HCQ increases pH in acidic organelles and has been reported to potently inhibit endosomal TLR activation. Whether HCQ can prevent endogenous TLR7 cleavage in primary immune cells, such as plasmacytoid dendritic cells (pDCs), had never been examined. Here, using a validated anti-TLR7 antibody suitable for biochemical detection of native TLR7 protein, we show that HCQ-treatment of fresh PBMCs, CAL-1 leukemic and primary human pDCs inhibits TLR7 cleavage and results in accumulation of full-length protein. As a consequence, we observe an inhibition of pDC activation in response to TLR7 stimulation with synthetic ligands and viruses including inactivated SARS-CoV2, which we show herein activates pDCs through TLR7-signaling. Together, our finding suggests that the major pathway by which HCQ inhibits ssRNA-sensing by pDCs may rely on its capacity to inhibit endosomal acidification and the functional maturation of TLR7 protein.

## Introduction

The Toll-like receptor 7 (TLR7) is essential for the induction of antiviral immunity but is also involved in the pathogenesis of systemic lupus erythematosus (SLE), an autoimmune disease with a strong female bias [1, 2]. TLR7 is a single-stranded RNA receptor encoded by an X-linked gene, and is maintained under strong purifying selection, which attests to non-redundant vital functions [3]. However, rare TLR7 genetic variants suppressing or modifying the function of the receptor have been recently identified in young male patients associated with severe COVID-19 [4].

TLR7 is expressed primarily in plasmacytoid dendritic cells (pDCs), B lymphocytes and monocytes [5, 6]. TLR7 protein exists in full size 140 kDa form in the endoplasmic reticulum (ER) and then undergoes post-ER proteolysis at the leucine-rich repeat (LRR) LRR14 and LRR15 regions, resulting in a C-term fragment of 75 kDa [7]. Crystallographic analysis of TLR7 shows that the C-term fragment remains associated with the N-term part of TLR7 to form a functional receptor in endosomes with two binding sites one for guanosine and another for ssRNA polyuridines [8].

Chloroquine (CQ) and hydroxychloroquine (HCQ) are used as first-line of treatment in patients with systemic lupus erythematosus (SLE) and rheumatoid arthritis, and have been shown to prevent endosomal TLR activation and type I IFN production by pDCs [9]. The ability of these drugs to inhibit activation of Toll-like receptors (TLR9, TLR8, and TLR7) has been attributed to their capacity to interact with nucleic acid ligands [10], rather than to the inhibition of endosome acidification, which is necessary for the maturation of these receptors [11, 12]. Conflicting results exist regarding the contribution of acid-dependent protease activity in the processing of endosomal TLR7 between mouse and human. Study in mouse cells have reported that processing of TLR7 and TLR9 does not take place in the absence of acidification of the endolysosomal compartments and requires stepwise processing with proteases [7, 13]. By contrast, processing of human TLR7 has been reported to be mediated by furin-like proteases at neutral pH, suggesting differential requirement for TLR7 processing between human and mouse [14]. So far, assessment of the effect of inhibition of endosomal acidification by HCQ or bafilomycin A1 on endogenous TLR7 cleavage in primary human immune cells has not been examined due to the lack of suitable antibodies for biochemical detection of TLR7.

In the present study, we investigated the impact of HCQ on the processing of human endogenous TLR7 in whole PBMCs, in the leukemic pDC line CAL-1, as well as in primary human pDCs using a highly-specific anti-TLR7 antibody [6, 15]. We also measured the half-life of TLR7 and the impact of HCQ on pDC activation in response to synthetic TLR ligands, as well as viruses, including SARS-CoV2.

## Results and Discussion

### Inhibition of endosomal acidification in fresh PBMCs inhibits TLR7 cleavage and results in accumulation of full-length protein

Analysis of human TLR7 protein expression in primary cells has been hampered due to the lack of suitable specific antibodies. We recently validated a commercially available monoclonal antibody directed against the C-terminus of human TLR7 that correctly detected both the full-length (140 kDa) and proteolytically mature (75 kDa) forms of natural and recombinant TLR7 in immunoblot analysis of human PBMCs [6, 15] (Fig. 1A-B). We investigated the impact of 18 hour-HCQ exposure of fresh PBMCs on TLR7 protein cleavage. We observed by immunoblot a dose dependent inhibition in the accumulation of the truncated 75 kDa C-term form of TLR7, which was concomitantly associated with an enhanced expression of the full-length 140 kDa TLR7 protein (Fig. 1C). As a consequence, the ratio between the full length versus truncated form of TLR7 was reduced for HCQ concentrations from 5 to 15 µM. As expected, blockade of autophagy by HCQ resulted in accumulation of LC3-II (Fig. 1D). Interestingly, similar results were observed using bafilomycin A1, an inhibitor of the vacuolar H^+^-ATPase which blocks acid-dependent protease activity (Fig. 1E-G). Overnight incubation of fresh PBMCs with bafilomycin A1 at low doses ranging from 50 to 100 nM inhibited by more than 70% the expression of the cleaved 75 kDa form of human TLR7 (Fig. 1E-G). Thus, endosomal acidification is critical for processing of endogenous TLR7 in human PBMCs.

**Figure 1.**
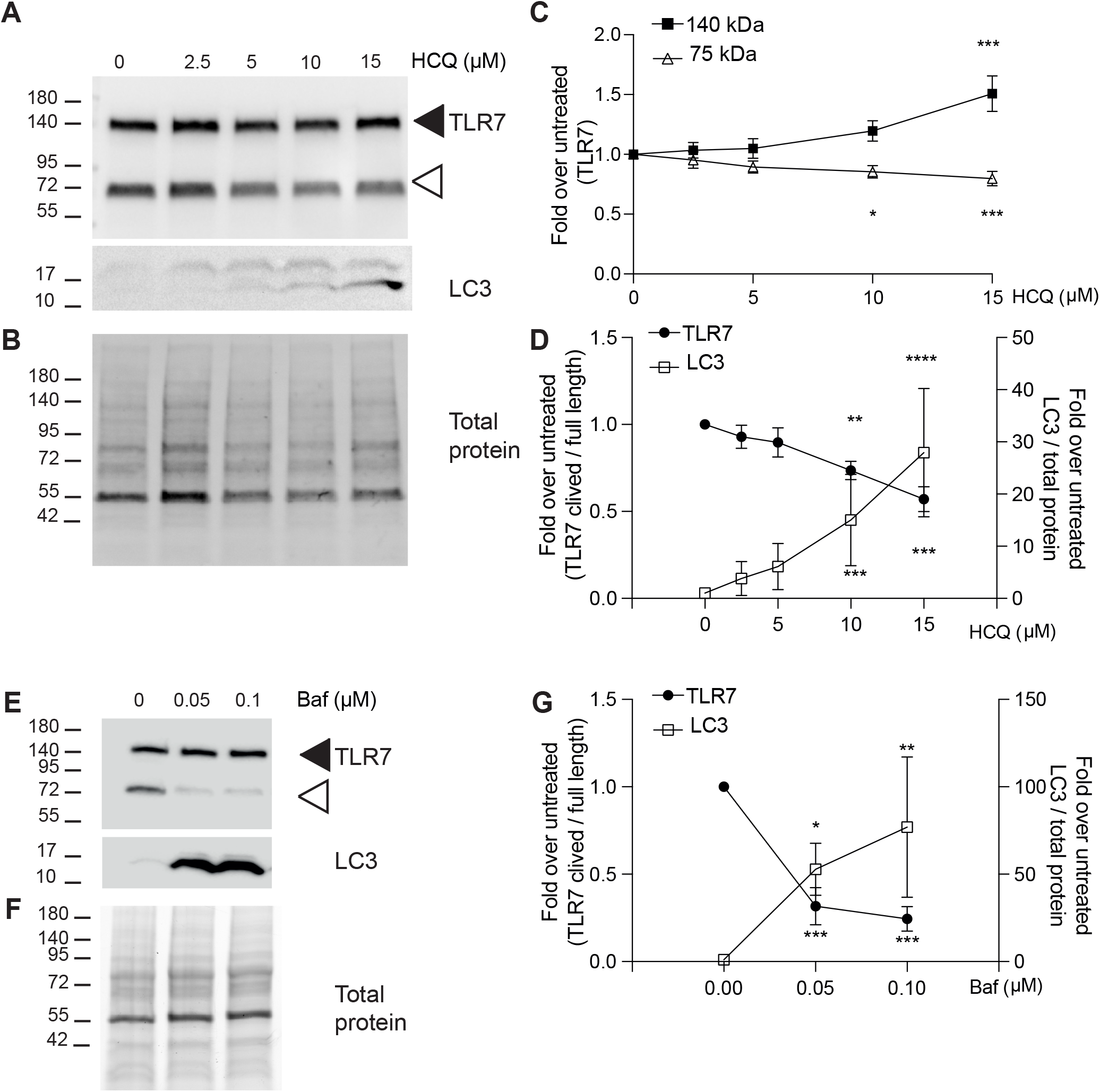
HCQ and bafilomycin A1 inhibit processing of TLR7 in human PBMCs. Freshly isolated PBMCs were incubated with various doses of HCQ (A – D) or bafilomycin A1 (Baf) (E – G) for 18 hours. 1×10^6^ cells were analyzed by immunoblot for TLR7 and LC3 (A, E) and total protein (B, F). Black arrowheads indicate 140 kDa full-length TLR7, white arrowheads indicate the 75 kDa proteolytically mature form of TLR7. (C, G) Densitometric quantification of TLR7. Band intensity is expressed as a ratio of treated over untreated after normalizing for total protein. (D) Densitometric analysis of TLR7 expressed as a ratio of TLR7 mature over full-length. For densitometric quantification of LC3-II, band intensity was normalized to total protein and expressed as a fold-change compared to untreated cultures. Data are expressed as mean ± SEM of ten different donors and are combined from two experiments for HCQ analysis and four different donors from one experiment for Baf analysis. Statistical significance was analyzed using two-way ANOVA followed by Dunnett test. Significance is indicated by *(p<0.05), **(p<0.01), ***(p<0.001), or ****(p<0.0001).

### HCQ inhibits TLR7 processing in human leukemic and primary pDCs

We first estimated the half-life of TLR7 protein in the leukemic CAL-1 pDC cell line. CAL-1 cells were exposed to cycloheximide (CHX) and then collected at different time points to quantify the proportion of the full-length and the processed forms of TLR7 relative to total protein levels (Fig. 2A-B). CHX-treatment resulted in a rapid decline in the relative proportion of both 140 kDa and 75 kDa TLR7 proteins in CAL-1 cells. From these results, we estimated the half-life of TLR7 protein to be in the range of 4 to 6 hours. These results indicate that TLR7 mRNA is constitutively translated into full-length TLR7 protein in CAL-1 cells probably as a consequence of constitutive expression of TLR7 mRNA transcripts in pDCs at single cell resolution [15, 16]. These results also show that the functionally active processed form of TLR7 follows the same kinetics, suggesting that continuous processing of TLR7 is required for its accumulation and activity in endosomes [14].

**Figure 2.**
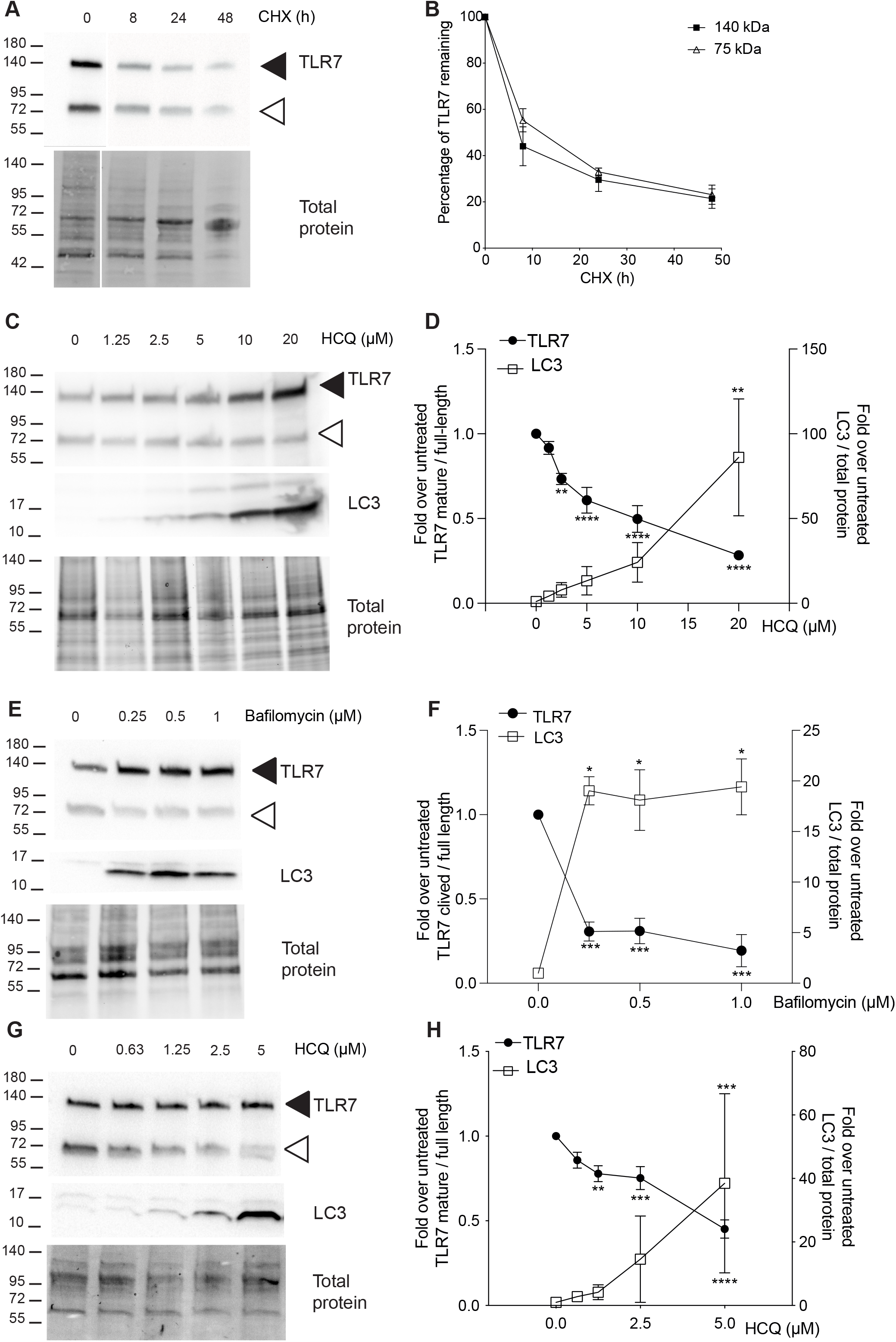
HCQ and Bafilomycin A1 inhibit the cleavage of endogenous TLR7 in human pDCs. (A, B) Measurement of TLR7 half-life by cycloheximide chase assay. CAL-1 were treated with cycloheximide (final concentration 50 µg/mL) for the indicated times and immunoblot was performed for TLR7. 1.5×10^5^ cells were loaded per lane. (A) TLR7 immunoblot and total protein representative of 3 independent experiments. Black arrowheads indicate 140 kDa full-length TLR7, white arrowheads indicate the 75 kDa proteolytically mature form of TLR7. (B) Densitometric quantification of the TLR7 protein forms was normalized to total protein and expressed as percentage of the corresponding band in the untreated (i.e., without cycloheximide) condition. Data are expressed as mean ± SEM (n=3 independent experiments). (C-F) Effect of HCQ (C, D) and Baf (E, F) on TLR7 cleavage in CAL-1 leukemic pDC treated with various doses of HCQ or Baf for 18h. 1.5×10^5^ cells were loaded per lane and immunoblotted for TLR7 and LC3. Black arrowheads indicate 140 kDa full-length TLR7, white arrowheads indicate the 75 kDa proteolytically mature form of TLR7. (D, F) Densitometric analysis of TLR7 expressed as a ratio of TLR7 mature over full-length and expressed as a fold change compared to untreated cells. For densitometric quantification of LC3-II, band intensity was normalized to total protein and expressed as a fold-change compared to untreated cultures. (G, H) Effect of HCQ on primary IL-3-pDCs treated with various doses of HCQ for 18h and immunoblotted for TLR7 and LC3. Results are expressed as mean ± SEM of three to five independent experiments (for CAL-1) or ten different donors combined from five experiments (for pDCs). Statistical significance was analyzed using two-way ANOVA followed by Dunnett test. Significance is indicated by *(p<0.05), **(p<0.01), ***(p<0.001), or ****(p<0.0001).

We next investigated the effect of 18 hour-HCQ treatment on the cleavage of TLR7 protein in CAL-1 cells. Pre-treatment of CAL-1 cells with HCQ doses ranging from 20 to 2.5 µM of HCQ resulted in a significant inhibition in the generation of the truncated TLR7 75 KDa form (Fig. 2C-D). As expected, blockade of autophagy by HCQ resulted in accumulation of LC3-II in CAL-1 cells (Fig. 2C-D).

To confirm that TLR7 processing occurs in an acidic environment in human pDCs, we measured the impact of bafilomycin A1 on TLR7 cleavage in CAL-1 cells. Bafilomycin A1 has been reported to prevent TLR9 receptor proteolysis, resulting in accumulation of full-length protein [7]. We show that 250 nM of bafilomycin leads to an almost complete inhibition of TLR7 cleavage which correlated to an accumulation of LC3-II (Fig. 2E-F), and was detected as soon as 2 to 4 hours of incubation with either HCQ or bafilomycin A1 (Supporting information Fig. S1). These results suggest that processing of endogenous TLR7 is a continuous process occurring in the endosomal acidic compartment in the leukemic CAL-1 pDC cell line, in agreement with published results regarding TLR9 processing [7].

To assess the impact of HCQ on endogenous TLR7 cleavage in primary human pDCs, we used an *in vitro* assay where human primary pDCs were cultured in the presence of IL-3 [17]. As for the CAL-1 cells, we found that 18 hour-treatment of primary IL-3-pDCs with HCQ resulted in significant inhibition of TLR7 processing at concentrations ranging from 5 to 1.25 µM of HCQ (Fig. 2G-H). Again, we observed a strong correlation between accumulation of LC3-II in HCQ-treated IL-3-pDCs and the dose-dependent inhibition in the accumulation of the C-term 75 KDa truncated form of TLR7 (Fig. 2G-H). Together, these results show that in leukemic CAL-1 cells and in primary human pDCs, HCQ can prevent the proteolytic cleavage of TLR7 protein.

### HCQ diminishes TLR7 signaling pathway in CAL-1 cells

Because it is well established that TLR7 processing is required for functional activity, we evaluated the impact of HCQ treatment on proximal signaling events in CAL-1 cells stimulated with imiquimod. Immunoblots were used to evaluate the phosphorylation of IRAK1, Akt and p38 as well as the degradation of IκBα (Supporting information Fig. S2A). IRAK1 phosphorylation by IRAK4 recruited to Myd88 is an early event of TLR7-activation leading to IRF7 activation and type I IFN production in pDCs [18]. As early as 30 minutes after TLR7 stimulation we observed a defect in IRAK1 phosphorylation in HCQ treated cells compared to untreated cells, which was maintained at 60 minutes (Supporting information Fig. S2A-B). We observed a similar profile for the imiquimod-induced phosphorylation of Akt and p38-MAPK (Supporting information Fig. S2C-D, respectively). Moreover, altered degradation of IκBα after HCQ treatment was also observed pointing to a general defect in TLR7-signaling in HCQ-treated cells (Supporting information Fig. S2E). At steady state, expression of these downstream signaling proteins, including IRF7, was not affected by HCQ-treatment nor cell-viability (Supporting information Fig. S2 and Fig. S3).

### Activation of pDCs by SARS-CoV2 is mediated through TLR7 and is blunted by HCQ treatment

As HCQ downregulates TLR7-dependent proximal signaling, we analyzed the impact of HCQ treatment on the capacity of IL-3-pDCs to produce IFN-α, IP-10 and TNF-α in response to activation with TLR7 ligands, including viruses. As in Fig. 2G, IL3-pDCs were incubated for the last 18 hours with various concentrations of HCQ before assessment of their functional responses. As shown in Fig. 3 and Supporting information Fig. S4, an almost complete inhibition of cytokine production was found for all doses of HCQ tested including the lowest one upon stimulation with the TLR7/8 ligand CL097. A similar trend was observed when pDCs were stimulated with betapropiolactone (BPL)-inactivated influenza A/Puerto Rico/8/1934(H1N1) virus (PR8) and BPL-inactivated Occitanie/2020/SCoV2-006 (SARS-CoV2) (Fig. 3 B-C). Likewise, the response of IL-3-pDCs stimulated through TLR9 with CpG-C was also totally inhibited upon HCQ treatment (Supporting information Fig. S4G). This inhibitory action of HCQ was not associated with enhanced cell death or apoptosis within the dose-range tested, and HCQ-treated IL-3-pDCs were still able to produce large amounts of TNF-α when activated with PMA and ionomycin (Supporting information Fig. S4H-I).

**Figure 3.**
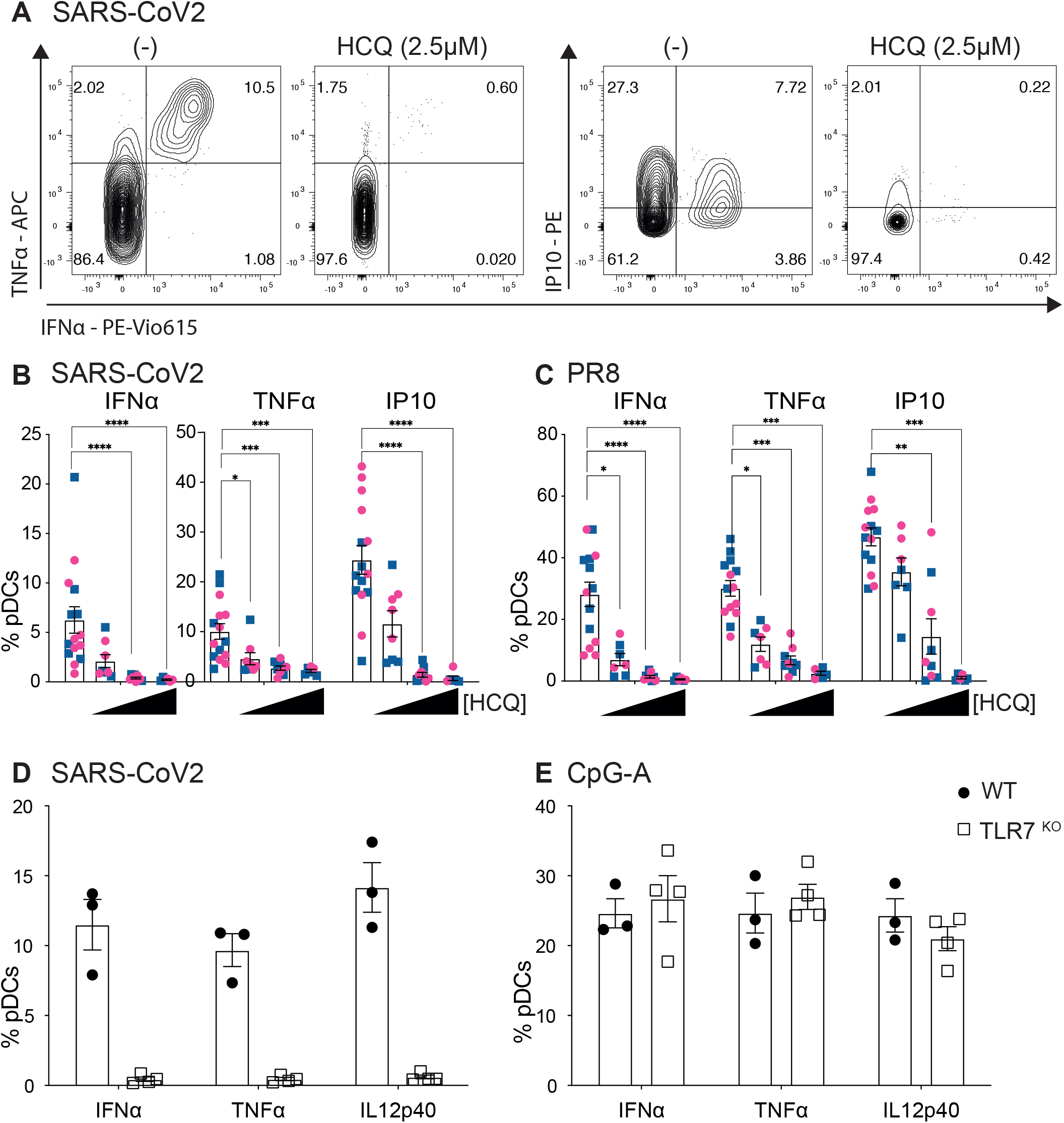
HCQ inhibits TLR7-dependent recognition of SARS-CoV2 by pDCs. pDCs were enriched from frozen PBMCs. After 2 days of culture with IL-3 (5 ng/mL), HCQ was added for 18 hours at different concentrations. Cells were then stimulated with the indicated TLR ligands or BPL-inactivated viruses for 4 hours. (A) Representative dot plots illustrating the intracellular cytokine staining of pDCs stimulated with SARS-CoV2, untreated or treated with 2.5 µM of HCQ. (B, C) Analysis of IFNα, TNFα and IP10-producing pDCs treated with increasing doses of HCQ from 1.25 to 5 µM. Histograms show the frequency of pDCs expressing the indicated cytokines after stimulation with SARS-CoV2 (B), PR8 (C). Men are represented with blue squares and women with pink circles. Data are expressed as mean ± SEM of 8 to 14 different donors and are combined from five experiments. Statistical significance was analyzed using two-way ANOVA followed by Dunnett test. Significance is indicated by *(p<0.05), **(p<0.01), ***(p<0.001), or ****(p<0.0001). (D, E) Bone marrow pDCs from wild type (black circles) or TLR7-deficient mice (open squares) were stimulated with inactivated SARS-CoV2 (D) or CpG-A (E). Histograms show the frequency of pDCs expressing the indicated cytokines after 4 hours of stimulation. Data are expressed as mean ± SEM of 3 to 4 different mice.

Our results confirmed recent observations that SARS-CoV2-induced pDC activation can be inhibited by HCQ [19]. Using genetically deficient patients, it was reported that pDC activation was critically dependent on IRAK4 and UNC93B1 [19]. However, because these genes can control the endosomal responses mediated by TLR7, TLR8 and TLR9, the exact nature of the TLR involved in pDC activation by SARS-CoV2 was unclear. As SARS-CoV2 is a ssRNA virus, we assess the role of TLR7 using mice deficient for this TLR. As shown in Fig. 3D and Supporting information Fig. S4, BPL-inactivated SARS-CoV2 readily activates the production of IFN-α, IL-12p40 or TNF-α in bone marrow pDCs from wild type but not in pDCs from TLR7-deficient mice. As a control, the TLR9-dependent response to CpG-A stimulation was preserved in TLR7-deficient animals (Fig. 3E, Supporting information Fig. S4). Thus, in response to inactivated SARS-CoV2, activation of pDC innate function is exclusively mediated through TLR7 signaling.

## Concluding remarks

This study reveals that the half-life of TLR7 protein is relatively short in human pDCs and requires continuous translation to maintain suitable processed TLR7 receptor for functional activation. Because of this short half-life, blockade of TLR7 processing with the drug HCQ or bafilomycin A1 rapidly results in the reduction in the functionally active form of the protein in the CAL-1 pDC cell line, but also in primary human pDCs. We now provide strong evidence that acid-dependent protease activity is critical for endogenous TLR7 processing in human pDCs, contrary to previous observations in TLR7-transduced THP-1 monocyte cell line [14]. Of note, cleavage inhibition of endogenous TLR7 was also evident in fresh PBMCs after 18 hours incubation with HCQ or bafilomycin A1 suggesting that acid-dependent endosomal proteases are critical for endogenous TLR7 processing across various cell types, including monocytes and B cells. Indeed, deconvolution experiments regarding the source of TLR7 expression by human PBMCs revealed that monocytes, pDCs and B cells were the main contributors of TLR7 protein expression at steady state [6].

Our results add to the list of the off-target effects of HCQ as an anti-viral drug [9]. Early activation of TLR7-dependent innate immunity is likely critical to control SARS-CoV2 infection as revealed by the occurrence of severe COVID-19 in patients with inborn errors of immunity targeting the type I IFN [20] or TLR7 [4] pathways. We propose that the anti-viral properties of HCQ could be counterbalanced by its capacity to blunt protective immunity linked to TLR7 activation or other immunomodulatory mechanisms [9]. This may contribute to explain the lack of beneficial action of HCQ *in vivo* on COVID-19 [21, 22] or SARS-CoV2 infection in non-human primates [23]. Indeed, *in vivo* treatment with HCQ in SLE patients has been shown to inhibit TLR7 and TLR9-dependent activation of pDCs upon *ex vivo* exposure of pDCs with specific TLR ligands [24]. Moreover, SLE patients with long-term treatment with HCQ are not protected from severe COVID-19 [25].

Our finding suggests that the major pathway by which HCQ inhibits TLR7-signaling may rely on its capacity to inhibit endosomal pH acidification thereby blunting the functional maturation of TLR7 protein. However, we cannot rule out that inhibition of interactions between TLR and nucleic acids [10] could be an additional mechanism of action of HCQ, particularly for TLR9, which has been reported to be more sensitive than TLR7 pathway for HCQ-mediated inhibition of pDC innate function *in vivo* [24]. Whether HCQ protection in SLE is associated with inhibition of proteolytic cleavage of endosomal TLRs will warrant further investigations.

## Materials and methods

### Cell culture, preparation, and stimulation

The CAL-1 human pDC cell line [26] was grown in R10 media which is: complete RPMI 1640 medium supplemented with 2 mM L-Glutamine, 1 mM sodium pyruvate, 100 U/mL penicillin/streptomycin, non-essential ammino acids, 50 µM 2-ME (all from Invitrogen) to which 10 % heat inactivated fetal bovine serum (Sigma) was added. Cells were cultured at 37°C in a CO_2_ in air incubator. Cells were seeded into 24-well plates (1 × 10^6^ cells / well) and incubated for 18 hours with hydroxychloroquine or bafilomycin A1 (Sigma). CAL-1 were stimulated with R837 (5 µg/mL) (Invivogen) for 30 or 60 minutes. PBMCs were freshly isolated from whole blood by Pancoll (PAN-Biotech) gradient separation and resuspended in R10 media. Cells were seeded in 48-well plates (5×10^5^ cells / well) and incubated for 18 hours with HCQ. All blood samples came from anonymous healthy donors from Etablissement Français du sang (Toulouse, France) in compliance with French regulations. Human pDCs were isolated from total frozen PBMC (obtained from a biobank authorized under file number 2-15-36 by the competent ethics board (Comité de Protection des Personnes Sud-Ouest et Outre-Mer II, Toulouse), using EasySep Human Plasmacytoid DC Isolation Kit (#17977 Stemcell) according to manufacturer protocol. Cells were seeded in 96-well plates (3×10^5^ cells / well), incubated for 48h with IL-3 (5 ng/mL) (Peprotech) and the last 18 hours with HCQ. Cells were then stimulated with inactivated SARS-CoV2 (25 μg/mL), PR8 (40 μg/mL), CL097 (50 ng/mL), Class C CpG ODN 2395 (2.5 µg/mL) (Invivogen) or PMA/ionomycin (50 and 500 ng/ml, respectively) for 4 hours with Brefeldin A (eBioscience) for the last 2 hours.

### Virus preparation and inactivation

PR8 was grown on Madin-Darby Canine Kidney (MDCK) cells (ATCC) in DMEM (Dutscher) supplemented with 100 U/mL penicillin, 100 μg/ml streptomycin (Invitrogen), and 1 µg/ml TPCK trypsin. SARS-CoV2 was grown on Vero E6 cells (ATCC) in DMEM (Dutscher) supplemented with 100 U/mL penicillin, 100 μg/ml streptomycin (Invitrogen), and 2% heat inactivated fetal bovine serum (Sigma). Viral stocks were propagated in 300 cm^2^ flasks (Dutscher) on which 10^3^ tissue culture infectious dose 50 (TCID_50_) of the respective viruses were inoculated in 100 ml infection media for 3 days at 37°C with 5% CO_2_. Each virus (culture supernatant) was harvested and inactivated with BPL (Fischer) overnight at 4°C. Each virus was concentrated by ultracentrifugation. Briefly, viral supernatant was transferred in Ultra-Clear centrifuge tubes (Beckman) on a 20% sucrose cushion. Then they were centrifuged at 4°C for 2 hours at 25 000 rpm using the Beckman SW-32 Ti rotor. Pellets were resuspended in PBS and stored at -80°C until use.

### Mouse cell stimulation

Male mice used were 18 weeks-old C57BL/6J (WT) or B6.129S1-Tlr7^tm1Flv^/J (Jackson Laboratory). Animals were housed in specific pathogen-free conditions. All mice were handled according to the Animal Care and Use of Laboratory guidelines of the French Ministry of Research. Bone marrow cells were isolated by flushing femurs with RPMI. Cells were seeded in 24-well plates (5 × 10^6^ cells / well) and stimulated with SARS-CoV2 (20 μg/mL) or Class A CpG, ODN 1585 (20 µg/mL) (Invivogen) for 4 hours with Brefeldin A for the last 2 hours.

### Western blotting

Cell lysates were prepared in Laemmli sample buffer (Invitrogen) and sonicated for 10 seconds. Total protein was quantified by BCA Protein Assay Kit (Pierce). Samples were heated for 10 minutes at 70°C in the presence of reducing agent (Invitrogen). Equal amounts of protein were subjected to SDS-PAGE on precast 4%–15% gradient gels (Bio-Rad). Gels were activated by ultraviolet (UV) exposure using a Bio-Rad ChemiDoc MP imager. Proteins were then transferred to Amersham Hybond 0.45-μm PVDF membranes (GE Healthcare). The membranes were blocked with 5% skim milk and 0.1% Tween-20 in TBS, probed overnight with human TLR7 (rabbit monoclonal IgG, clone EPR2088[2]; Abcam) antibody and incubated with anti–rabbit IgG (#7074, Cell Signaling Technology). For signaling pathway, membranes were blocked with 5% BSA and 0.1% Tween-20 in TBS, probed with Phospho-IRAK (Thr209) (#12756) or Phospho-Akt (Ser473) (#4060) or Phospho-p38 MAPK (Thr180/Tyr182) (#4511) or IκBα (#4814) and incubated with anti-rabbit IgG or anti-mouse IgG (#7076) all from Cell Signaling Technology. The membranes were then imaged for Stain-Free staining and total protein was quantified using ImageLab 5.0 software (Bio-Rad). Chemiluminescent detection was carried out with Amersham ECL Select or ECL Prime reagents (GE Healthcare) as necessary.

### Flow cytometry

Cells were stained for viability with Viability Dye-eFluor506 (eBiosciences). For human’s surface markers, cells were stained in FACS solution with the following antibodies: BDCA4-PB (clone 12C2, Biolegend), and HLA-DR-APC Vio770 (clone REA805, Miltenyi Biotech). Cells were fixed with fixation/permeabilization solution (BD Biosciences). For intracellular staining cell were permeabilized in Perm / wash solution (BD Biosciences) and stained with: IFN-α-PE Vio615 (clone REA1013, Miltenyi Biotec), TNF-α-APC (clone Mab11, Biolegend), and IP-10-PE (clone J034D6, Biolegend). Mouse cells were stained with the following antibodies: PDCA1-PE-Cyanine7 (clone eBio927, eBiosciences) and B220-APC-Cyanine7 (clone RA3-6B2, Invitrogen). For intracellular staining, cell were permeabilized in Perm/wash solution (BD Biosciences) and stained with : anti-IFNα-APC (clone RMMA-1, Invitrogen), anti-TNFα-PE (clone MP6-XT22, BD Biosciences), and anti-IL-12p40-PE-Cy5.5 (clone C17.8, Invitrogen). Samples were acquired on LSR II or Fortessa (BD Biosciences). The gating strategy is displayed as Supporting Information Fig. S4 and Fig. S5.

### Data analysis and statistics

FACS analyses were made by using FlowJo software v10 (TreeStar). Statistical analyses were carried out using GraphPad Prism 8 software (GraphPad). Statistical significance was analyzed using two-way ANOVA followed by Dunnett test.

## Supporting information

Supporting Informations

## Data Availability

The data that support the findings of this study are presented in this paper. No shared databases were used or created.

## ACKNOWLEDGMENTS

We are indebted to the core facilities at INFINITY INSERM U1291 and at INSERM US006. This work was supported by grants from Agence National de la Recherche (ANR-20-COV8-0004-01 and ANR-20-CE15-0014-01), the FOREUM Foundation and the Occitanie Region (INSPIRE research program).

## AUTHOR CONTRIBUTIONS

CC and JCG conceived and designed the study. CC performed experiments and analyzed the data. MD contributed essential reagents. JCG wrote the manuscript with input from the co-authors. All authors read and approved the final version of the manuscript.

## List of abbreviations

(TLR): Toll-like receptor
(HCQ): Hydroxychloroquine
(pDCs): plasmacytoid dendritic cells
(SLE): Systemic lupus erythematosus
(BPL): betapropiolactone

