## Supporting Informations for "Hydroxychloroquine inhibits proteolytic processing of endogenous TLR7 protein in human primary plasmacytoid dendritic cells"

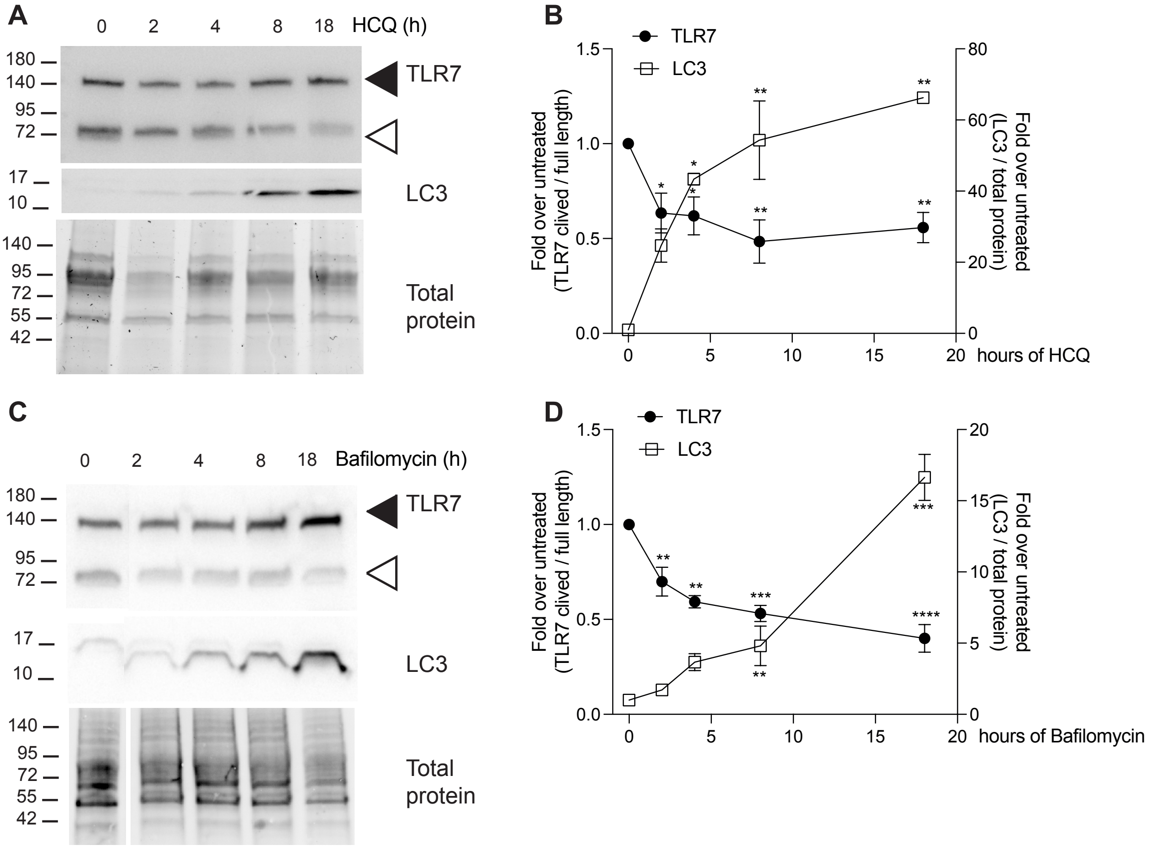


**Supporting information Figure S1. HCQ and Bafilomycin inhibit processing of TLR7 in a time dependent manner.** CAL-1 cells were incubated for indicated times with HCQ (10 µM) (A, B) or Baf (0.5 µM) (C, D). 1 x $10^5$ cells were analyzed for TLR7 and LC3 by immunoblot (A, C). Black arrowheads indicate 140 kDa full-length TLR7, white arrowheads indicate the 75 kDa proteolytically mature form of TLR7. (B, D) Densitometric quantification of TLR7 and LC3. Densitometric analysis of TLR7 and LC3 as in Fig. 1. Data are expressed as mean ± SEM of 3 independent experiments. Statistical significance was analyzed using two-way ANOVA followed by Dunnett test. Significance is indicated by *(p<0.05), **(p<0.01), ***(p<0.001), or ****(p<0.0001).

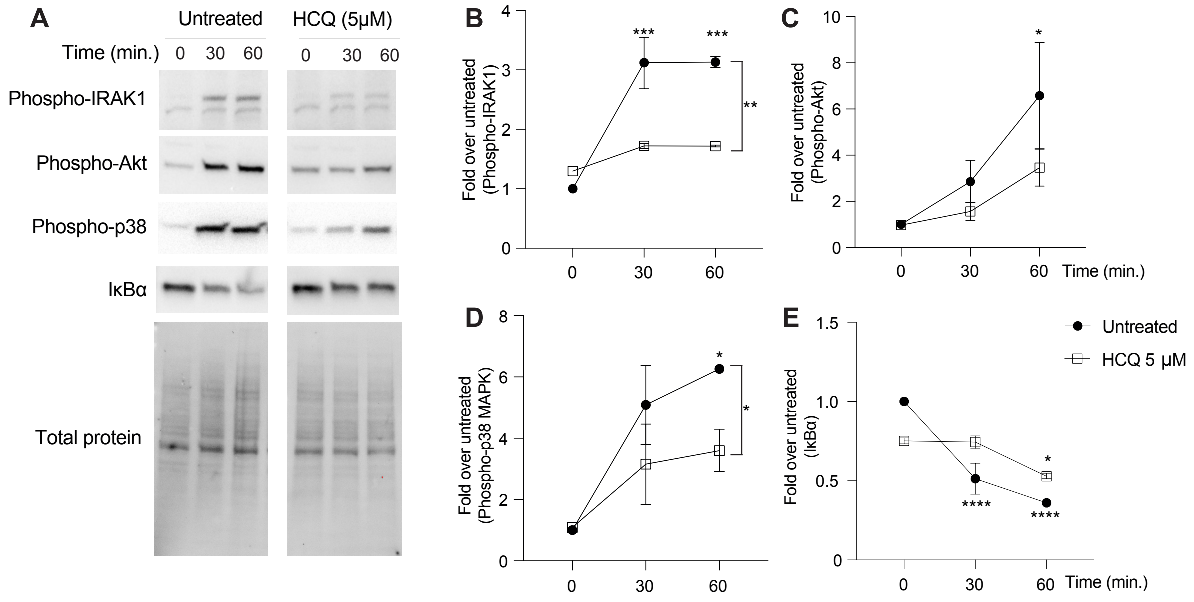


**Supporting information Figure S2. HCQ inhibits TLR7-dependent proximal signaling.** CAL-1 were pre-treated with 5µM HCQ for 18 hours and stimulated with R837 for the indicated times and immunoblots were performed on 1 x $10^5$ cells for P-IRAK1, P-Akt, P-p38 and IκBα (A). (B-E) Densitometric quantification of P-IRAK1 (B), P-Akt (C), P-p38 (D) and IκBα (E) normalized to total protein and expressed as a ratio compared to untreated, unstimulated cells. Results are expressed as mean ± SEM of three independent experiments. Statistical significance was analyzed using two-way ANOVA followed by Dunnett test. Significance is indicated by *($p<0.05$), **($p<0.01$).

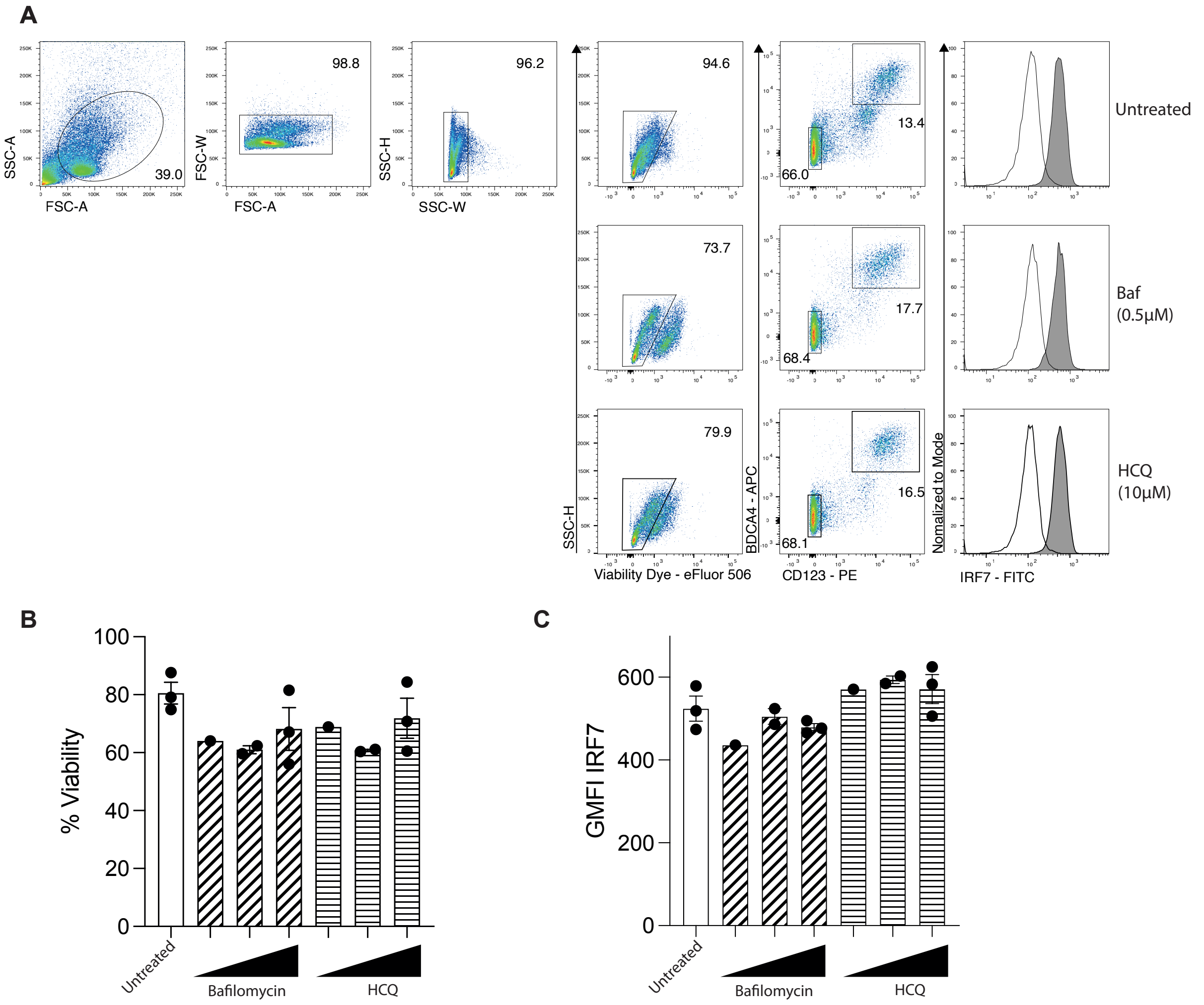


**Supporting information Figure S3. HCQ or bafilomycin neither alter cell biability nor IRF7 expression in primary human pDCs.** Human pDCs were purified from frozen PBMCs. After 2 days of culture with IL-3 (5 ng/mL), HCQ or Baf were added for 18 hours at different concentrations. Cells were analyzed for their viability and for IRF7. (A) Representative gating strategy for pDCs staining. Histograms show the frequency of live pDCs (B) and IRF7 GMFI upon pDCs (C).

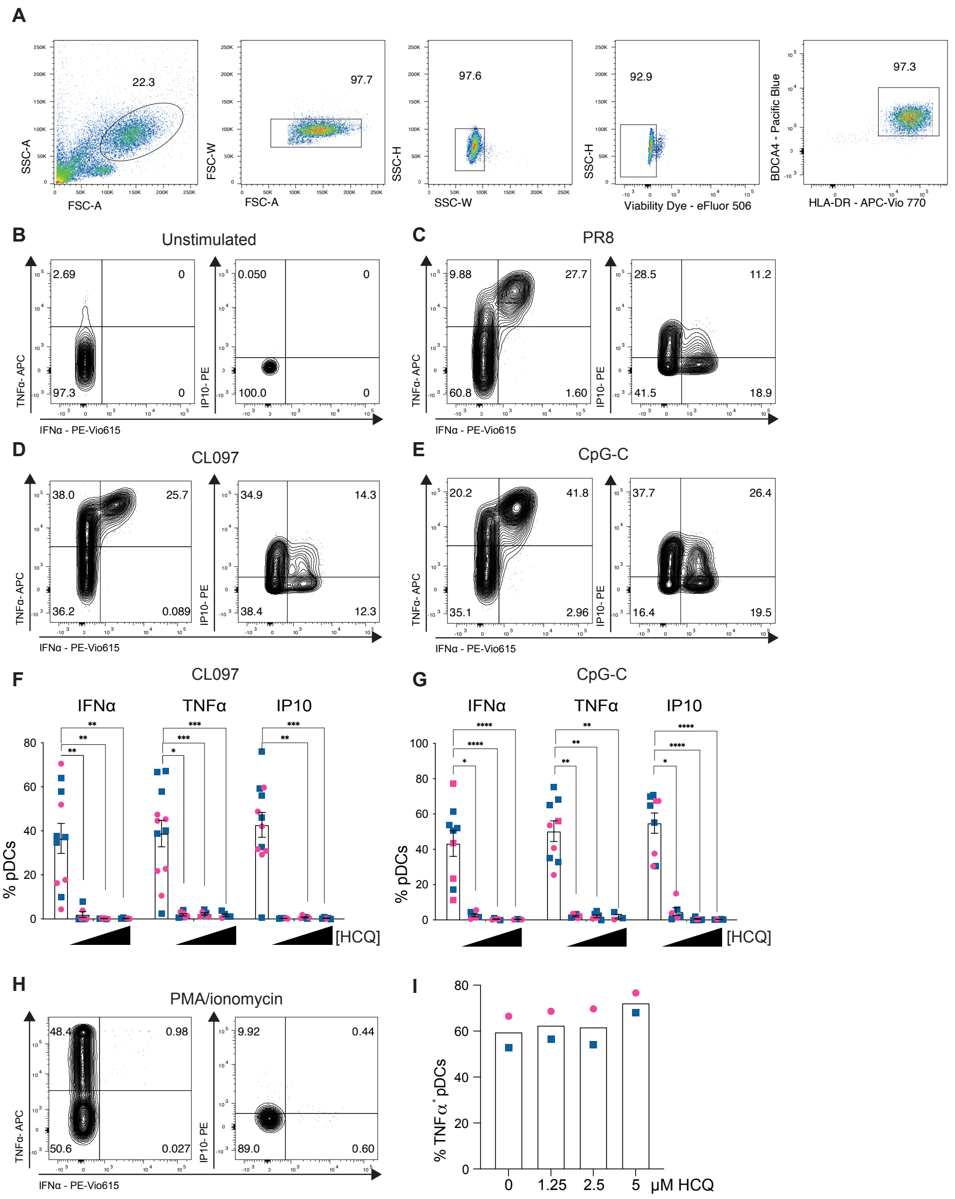


**Supporting information Figure S4. HCQ inhibits the functional responses of pDCs stimulated through either TLR7 or TLR9.** pDCs were purified from frozen PBMCs. After 2 days of culture with IL-3 (5 ng/mL), HCQ was added for 18 hours at different concentrations. Cells were then stimulated with the indicated TLR ligands or BPL-inactivated viruses for 4 hours. (A) Representative gating strategy for pDCs identification. (B-E) representative dot plots illustrating the intracellular IFNα / TNFα staining of pDCs unstimulated (B) or stimulated with PR8 (C), CL097 (D) or CpG-C (E). (F, G) Histograms show the frequency of pDCs expressing the indicated cytokines after stimulation with CL097 (F) or CpG-C (G), same as in Fig3. (H, I) pDCs, treated with various doses of HCQ were stimulated with PMA/ionomycin. (H) Representative dot plot illustrating the intracellular IFNα / TNFα / IP10 staining of pDCs. (G) Histogram shows the frequency of pDCs positives for TNFα upon PMA/ionomycin stimulation.

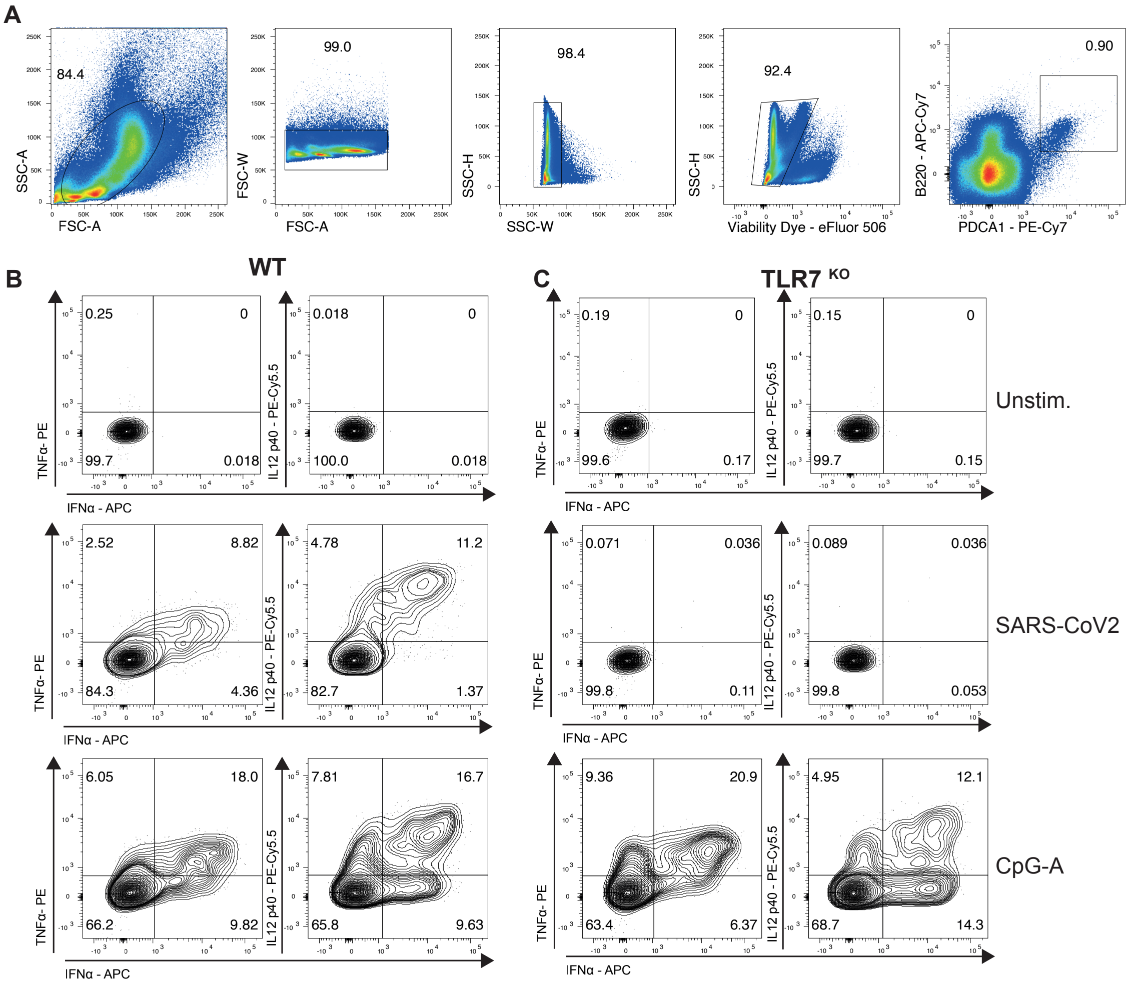


**Supporting information Figure S5. SARS-CoV2 activates mouse pDCs exclusively through TLR7.** (A) Representative gating strategy for bone marrow mouse pDCs identification. (B, C) Representative dot plot illustrating the intracellular staining of pDCs from WT (B) or TLR7 deficient mice (C) stimulated with SARS-CoV2 or CpG-A.
